# Instability in longitudinal sleep duration predicts cognitive impairment in aged participants of the Seattle Longitudinal Study

**DOI:** 10.1101/2023.06.07.23291098

**Authors:** Samantha A. Keil, Abigail G. Schindler, Marie X. Wang, Juan Piantino, Lisa C. Silbert, Jonathan E. Elliott, Ronald G. Thomas, Sherry Willis, Miranda M. Lim, Jeffrey J. Iliff

## Abstract

**Importance:** Sleep disturbances and clinical sleep disorders are associated with all-cause dementia and neurodegenerative conditions. It remains unclear how longitudinal changes in sleep impact the incidence of cognitive impairment.

**Objective:** To evaluate how longitudinal sleep patterns contribute to age-related changes in cognitive function in healthy adults.

**Design, Setting, Participants:** This study utilizes retrospective longitudinal analyses of a community-based study within Seattle, evaluating self-reported sleep (1993-2012) and cognitive performance (1997-2020) in aged adults.

**Main Outcomes and Measures:** The main outcome is cognitive impairment as defined by sub-threshold performance on 2 of 4 neuropsychological batteries: Mini-Mental State Examination (MMSE), Mattis Dementia Rating Scale, Trail Making Test, and Wechsler Adult Intelligent Scale (Revised). Sleep duration was defined through self-report of ‘average nightly sleep duration over the last week’ and assessed longitudinally. Median sleep duration, change in sleep duration (slope), variability in sleep duration (standard deviation, Sleep Variability), and sleep phenotype (“Short Sleep” median ≤7hrs.; “Medium Sleep” median = 7hrs; “Long Sleep” median ≥7hrs.).

**Results:** A total of 822 individuals (mean age of 76.2 years [11.8]; 466 women [56.7%]; 216 *APOE* allele positive [26.3%]) were included in the study. Analysis using a Cox Proportional Hazard Regression model (concordance 0.70) showed that increased Sleep Variability (95% CI [1.27,3.86]) was significantly associated with the incidence of cognitive impairment. Further analysis using linear regression prediction analysis (R^2^=0.201, F (10, 168)=6.010, p=2.67E-07) showed that high Sleep Variability (β=0.3491; p=0.048) was a significant predictor of cognitive impairment over a 10-year period.

**Conclusions and Relevance:** High variability in longitudinal sleep duration was significantly associated with the incidence of cognitive impairment and predictive of decline in cognitive performance ten years later. These data highlight that instability in longitudinal sleep duration may contribute to age-related cognitive decline.

## Introduction

Over 60 million people currently suffer from dementia globally, a number expected to more than double to 153 million over the next 30 years [1]. Alzheimer’s disease (AD) accounts for an estimated 60-80% of these dementia cases [2, 3]. Observational clinical studies demonstrate that many events precede the development of clinical cognitive decline, including amyloid β deposition that appears to begin at least 15 years before the onset of clinical cognitive impairment [4]. Data from interventional trials suggest that targeting pathogenic processes early in the course of disease may provide the most effective avenue for therapeutic intervention [5, 6]. Therefore, understanding the upstream processes contributing to the development of dementia in the decades preceding cognitive and functional decline may provide approaches to the treatment and prevention of dementing disorders.

Sleep disruption and dementia have long been associated in clinical populations. Emerging studies suggest that 40-90% of patients suffering from neurodegenerative conditions, including AD, suffer from disrupted sleep up to 30 years prior to the emergence of cardinal disease symptoms [7, 8]. Although this association was initially believed to reflect the progressive degeneration of sleep-regulatory centers and circuits throughout disease progression [9], more recent studies suggest that sleep disruption may contribute to the pathological processes underlying these dementing disorders. Short sleep duration has been associated with increased risk of cognitive impairment in healthy aging adults [10–13], and AD biomarker studies demonstrate that in cognitively intact study populations, poor sleep quality or short sleep duration are also associated with greater AD-related pathological burden [14, 15]. While these studies suggest a link between chronic sleep disruption, and dementia diagnosis, they are limited by their reliance on relatively simple, generally cross-sectional, measures of sleep quality and duration.

Here we evaluate the impact of longitudinal changes in sleep duration on age-associated cognitive decline within a community-based longitudinal study. Utilizing both longitudinal self-reported sleep duration and a cognitive battery regularly collected within the Seattle Longitudinal Study (SLS) [16], we first define the relationship between sleep duration, sleep quality, changes in sleep duration over time, and variability in sleep duration over time with the incidence of cognitive impairment in an aged population. We then assess whether these longitudinal patterns of sleep predict the development of cognitive impairment over a 10-year interval.

## Methods

### Sample

SLS participants were recruited from participants in the Group Health Cooperative of Puget Sound and Health Maintenance Organization of Washington and data was collected between 1956 and 2020 [16–20]. The present study evaluated a sub-sample within the SLS that were >55 years of age and underwent multiple rounds of both a mail-home Health Behavior Questionnaire (HBQ) and in-person or mail-home neuropsychological battery. Of the 1104 participants who received both the HBQ and neuropsychological battery, only participants with complete demographic information including *APOE* allele status were included in analysis, resulting in an initial study participant sample of n=822 (**Figure 1**).

**Figure 1.**
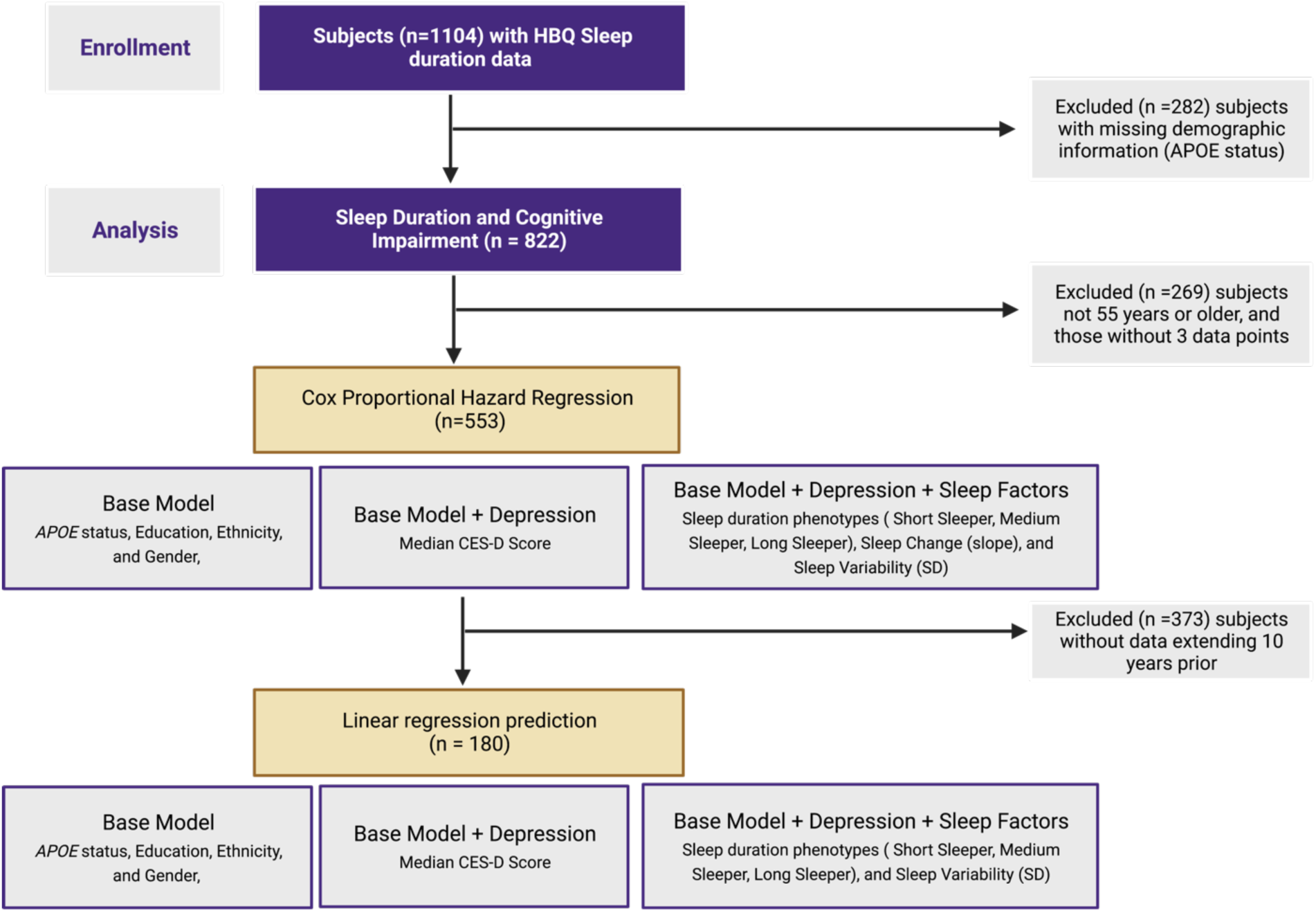
Flow chart Seattle Longitudinal Study participant inclusion and exclusion.

### Participant Testing

The HBQ was provided at 3–5-year intervals beginning in 1993 and contained a question asking: “In the PAST 7 DAYS, how many hours did you usually sleep per 24 hours (not counting naps)?”, with a Likert scale response of 1=5 hrs or less, 2=6 hrs, 3=7 hrs, 4=8 hrs, 5=9 hrs or more. From these longitudinal data, we derived multiple sleep measures for each participant: Median Sleep Duration (median value of all measurements), Change in Sleep Duration (best-fit slope of all measurements), Variability in Sleep Duration (standard deviation of all measurements), and Sleep Phenotype (classified as “Short Sleep”, <7 hrs; “Medium Sleep” =7 hrs; and “Long Sleep” >7 hrs).

The neuropsychological battery, performed every 5-7 years, was administered between 1997 to 2019 and included the Mini-Mental State Examination (MMSE)[21–23], Mattis Dementia Rating Scale (Mattis)[24–26], Trails Making Test B (Trails B)[27], and Wechsler Adult Intelligent Scale - Revised (WAIS) [28, 29]. Within the original SLS, participants whom performed below threshold on these tests, were deteremined cognitively impaired through consensus of two neuropsychologists [16–20]. In our analysis, in order to best capture subtle changes in cognitive impairment, we defined cognitive impairment based on performance below age-normed threshold on two out of four of the neuropsychological tests: MMSE score below 27, Mattis score below 130, Trails B score longer than 180 seconds, and age-adjusted scaled score less than 8 for WAIS Vocabulary, WAIS Comprehension, WAIS Block Design, and WAIS Digit Symbol. The SLS also included the the Center for Epidemiologic Studies–Depression Scale (CES-D) witin their neuropsychological testing battery [30, 31]. Designed to evaluate the frequency and severity of depressive symptoms through 20 likert scale questions, assessing a subcategory of symptoms including depressive affect, positive affect, somatic complaints (including sleep restlessness), and interpersonal difficulties. In this study, the median CES-D global score was used as a covariate throughout analysis.

### Statistical Analysis

All statistical analysis were completed with Python, all statistical tests were two-sided, and p-values of less than 0.05 were considered statistically significant. Baseline study characteristics for all demographic (gender, ethnicity, level of education, and Apolipoprotein Ɛ4 carrier status), depression (median CES-D total score), sleep measures (median self-reported sleep duration and sleep phenotype), and cognitive measures (MMSE, Mattis, Trails B, WAIS, and composite score) were evaluated binned by age groups of <65 yrs, 65-84 yrs, and ≥85 yrs. Statistical differences were assessed by χ^2^ test and One-Way ANOVA with Šídák’s multiple comparison test used for post hoc analysis when appropriate as noted in **Table 1**. Baseline characteristics were also performed on the participant sample utilized for Cox Proportional Hazard Regression and Linear Regression prediction analysis, to ensure that the samples were comparable.

**Table 1.** Baseline characteristics of study and sub study participants. SD: Standard Deviation; CES-D: Center for Epidemiological Studies Depression Scale; Mattis: Mattis dementia rating scale; WAIS: Wechsler Adult Intelligence Scale. †: 285 yrs. vs. <65 yrs.; ††: 285 yrs. vs. 65-84 yrs.; †††: 65-84 yrs. vs. <65 yrs. Šídák’s multiple comparison test was used for all Post Hoc analysis.

| Sample 1 (n=821) | All study participants |  |  |  |  |  |
| --- | --- | --- | --- | --- | --- | --- |
|  | <65 (n=150) | 65-84 (n=449) | ≥ 85 (n=223) | Statistical Test | p | Post Hoc Test p |
| **Age - Mean (SD) [Range]** | 57.59 (5.71) [40-64] | 75.72 (5.35) [65-84] | 89.78 (3.62) [85-100] |  |  |  |
| **Female Gender - Total (%)** | 87 (58.0) | 248 (55.2) | 131 (58.7) | $\chi^2$ | 0.6453 | |
| **Ethnicity - Total (%)** | | | | $\chi^2$ | 0.2267 | |
| White (Non-Hispanic) | 139 (92.7) | 429 (95.5) | 215 (96.4) |  |  |  |
| Non-White | 11 | 20 | 8 |  |  |  |
| **Education (%)** | | | | $\chi^2$ | 0.0003 | |
| Highschool or Below | 9 (6.0) | 69 (15.4) | 50 (22.4) |  |  |  |
| College | 74 (49.3) | 198 (44.1) | 103 (46.2) |  |  |  |
| Graduate School | 67 (44.7) | 182 (40.5) | 70 (31.4) |  |  |  |
| **Apolipoprotein E4 Carrier - Total (%)** | 35 (23.3) | 130 (29.0) | 51 (22.9) | $\chi^2$ | 0.1598 | |
| **Depression Measure** |  |  |  |  |  |  |
| **CES-D - Mean (SD) [Range]** |  |  |  |  |  |  |
| Total Score | 9.33 (7.16) [0-33] | 7.60 (5.84) [0-38] | 8.72 (5.62) [0-35] | One-Way ANOVA | 0.0037 | 0.0076<sup>†††</sup> |
| **Sleep Measures** |  |  |  |  |  |  |
| **Self-Reported Sleep Duration - Mean (SD)** |  |  |  |  |  |  |
| Sleep Duration | 6.65 (0.87) | 6.93 (0.87) | 7.06 (1.02) | One-Way ANOVA | <0.0001 | <0.0001<sup>†</sup>, 0.0035<sup>†††</sup> |
| **Sleep Phenotype - Total (%)** | | | | $\chi^2$ | 0.0004 | |
| Short Sleeper: Median <7 hrs. | 68 (45.3) | 145 (32.3) | 63 (28.2) |  |  |  |
| Medium Sleeper: Median 7 hrs. | 55 (36.7) | 168 (37.4) | 76 (34.1) |  |  |  |
| Long Sleeper: Median >7 hrs. | 27 (18.0) | 136 (30.3) | 84 (37.7) |  |  |  |
| **Cognitive Measures** |  |  |  |  |  |  |
| **Cognitive Test Score – Mean (SD) [range]** |  |  |  |  |  |  |
| Mini-Mental State Examination Score | 29.31 (1.19) [23-30] | 28.12 (2.42) [14-30] | 26.12 (4.16) [1-30] | One-Way ANOVA | <0.0001 | <0.0001<sup>†</sup>, <0.0001<sup>††</sup>, <0.0001<sup>†††</sup> |
| Mattis Dementia Rating Scale | 141.5 (2.57) [130-144] | 139.0 (5.85) [101-144] | 131.8 (14.79) [16-144] | One-Way ANOVA | <0.0001 | <0.0001<sup>†</sup>, <0.0001<sup>††</sup>, 0.009<sup>†††</sup> |
| Trail Making Test B | 62.17 (29.51) [25-285] | 113.5 (80.32) [19-689] | 173.5 (95.77) [4-579] | One-Way ANOVA | <0.0001 | <0.0001<sup>†</sup>, <0.0001<sup>††</sup>, <0.0001<sup>†††</sup> |
| WAIS - Vocabulary Age-Scaled | 14.67 (2.23) [6-19] | 13.53 (2.57) [4-19] | 12.32 (2.78) [2-19] | One-Way ANOVA | <0.0001 | <0.0001<sup>†</sup>, <0.0001<sup>††</sup>, <0.0001<sup>†††</sup> |
| WAIS - Comprehension Age-Scaled | 13.80 (2.55) [6-19] | 13.12 (2.62) [4-19] | 11.85 (2.69) [0-19] | One-Way ANOVA | <0.0001 | <0.0001<sup>†</sup>, <0.0001<sup>††</sup>, 0.0197<sup>†††</sup> |
| WAIS - Block Design Age-Scaled | 14.39 (2.74) [8-19] | 13.39 (2.99) [3-19] | 10.73 (3.30) [0-18] | One-Way ANOVA | <0.0001 | <0.0001<sup>†</sup>, <0.0001<sup>††</sup>, 0.014<sup>†††</sup> |
| WAIS - Digit-Symbol Age-Scaled | 12.78 (2.27) [7-19] | 12.57 (2.82) [1-19] | 9.65 (2.78) [0-16] | One-Way ANOVA | <0.0001 | <0.0001<sup>†</sup>, <0.0001<sup>††</sup> |
| Cognitively Impaired (%) | 10 (6.7) | 79 (17.6) | 107 (48.0) | $\chi^2$ | <0.0001 | |
| **Sample 2** |  |  |  |  |  |  |
| Participants evaluated with Cox Proportional Hazard Regression |  |  |  |  |  |  |
|  | < 65 (n=100) | 65-84 (n=314) | ≥ 85 (n=139) | Statistical Test | p | Post Hoc Test p |
| **Age - Mean (SD) [Range]** | 60.62 (2.57) [55-64] | 75.32 (5.55) [65-84] | 89.92 (3.57) [85-100] |  |  |  |
| **Female Gender - Total (%)** | 59 (59.0) | 175 (55.0) | 87 (64.4) | $\chi^2$ | 0.1743 | |
| **Ethnicity - Total (%)** | | | | $\chi^2$ | 0.0695 | |
| White (Non-Hispanic) | 91 (91.0) | 320 (96.4) | 127 (96.2) |  |  |  |
| Non-White | 9 | 12 | 5 |  |  |  |
| **Education - Total (%)** | | | | $\chi^2$ | 0.001 | |
| Highschool or Below | 4 (4.0) | 32 (10.2) | 25 (18.0) |  |  |  |
| College | 51 (51.0) | 133 (42.4) | 70 (50.4) |  |  |  |
| Graduate School | 45 (45.0) | 149 (47.5) | 44 (31.6) |  |  |  |
| **Apolipoprotein E4 Carrier - Total (%)** | 24 (24.0) | 96 (30.6) | 30 (21.6) | $\chi^2$ | 0.1032 | |
| **Depression Measure** |  |  |  |  |  |  |
| **CES-D - Mean (SD) [Range]** |  |  |  |  |  |  |
| Total Score | 9.43 (7.24) [0-29] | 6.99 (5.61) [0-38] | 8.27(5.58) [0-35] | One-Way Anova | 0.0008 | 0.0011<sup>†††</sup> |
| **Sleep Measures** |  |  |  |  |  |  |
| **Self-Reported Sleep Duration - Mean (SD)** |  |  |  |  |  |  |
| Sleep Duration | 6.60 (0.88) | 6.83 (0.86) | 7.00 (0.94) | One-Way Anova | 0.0018 | 0.0012<sup>†</sup> |
| Sleep Change (Slope) | -0.14 (0.38) | -0.13 (0.32) | -0.18 (0.35) | One-Way Anova | 0.3171 |  |
| Sleep Variability (SD) | 0.81 (0.36) | 0.73 (0.31) | 0.71 (0.34) | One-Way Anova | 0.0585 |  |
| **Sleep Phenotype - Total (%)** | | | | $\chi^2$ | 0.0079 | |
| Short Sleeper: Median <7 hrs. | 48 (48.0) | 110 (35.0) | 39 (28.1) |  |  |  |
| Medium Sleeper: Median 7 hrs. | 36 (36.0) | 127 (40.4) | 54 (38.8) |  |  |  |
| Long Sleeper: Median >7 hrs. | 16 (16.0) | 77 (24.5) | 46 (33.1) |  |  |  |
| **Cognitive Measures** |  |  |  |  |  |  |
| **Cognitive Test Score - Mean (SD) [Range]** |  |  |  |  |  |  |
| Mini-Mental State Examination Score | 29.2 (1.33) [23-30] | 28.37 (2.18) [14-30] | 26.21 (4.52) [1-30] | One-Way Anova | <0.0001 | <0.0001<sup>†</sup>, <0.0001<sup>††</sup>, 0.0243<sup>†††</sup> |
| Mattis Dementia Rating Scale | 141.1 (2.85) [130-144] | 139.9 (4.76) [112-144] | 132.2 (17.15) [16-144] | One-Way Anova | <0.0001 | <0.0001<sup>†</sup>, <0.0001<sup>††</sup> |
| Trail Making Test B | 63.1 (30.11) [25-285] | 99.1 (70.15) [19-689] | 158.7 (88.74) [4-500] | One-Way Anova | <0.0001 | <0.0001<sup>†</sup>, <0.0001<sup>††</sup>, <0.0001<sup>†††</sup> |
| WAIS - Vocabulary Age-Scaled | 14.75 (2.22) [6-19] | 13.79 (2.41) [7-19] | 12.48 (2.47) [2-19] | One-Way Anova | <0.0001 | <0.0001<sup>†</sup>, <0.0001<sup>††</sup>, 0.0015<sup>†††</sup> |
| WAIS - Comprehension Age-Scaled | 13.97 (2.63) [6-19] | 13.32 (2.41) [7-19] | 11.83 (2.60) [0-19] | One-Way Anova | <0.0001 | <0.0001<sup>†</sup>, <0.0001<sup>††</sup> |
| WAIS - Block Design Age-Scaled | 14.43 (2.90) [8-19] | 13.94 (2.89) [3-19] | 11.36 (3.22) [0-18] | One-Way Anova | <0.0001 | <0.0001<sup>†</sup>, <0.0001<sup>††</sup> |
| WAIS - Digit-Symbol Age-Scaled | 12.81 (2.26) [7-19] | 13.13 (2.78) [1-19] | 10.08 (2.85) [0-16] | One-Way Anova | <0.0001 | <0.0001<sup>†</sup>, <0.0001<sup>††</sup> |
| Cognitively Impaired (%) | 5 (5.0) | 37 (11.8) | 52 (37.4) | $\chi^2$ | <0.0001 | |
| **Sample 3** |  |  |  |  |  |  |
| Participants undergoing predictive modeling |  |  |  |  |  |  |
|  |  | **< 85 (n=108)** | **≥ 85 (n=72)** | **Statistical Test** | **p** |  |
| **Age - Mean (SD) [Range]** |  | 76.49 (5.58) [57-84] | 90.31 (3.71) [85-99] |  |  |  |
| **Female Gender - Total (%)** |  | 71 (65.7) | 37 (63.9) | Fisher's exact test | 0.8736 |  |
| **Ethnicity - Total (%)** |  |  |  | Fisher's exact test | >0.9999 |  |
| White (Non-Hispanic) |  | 106 (98.2) | 70 (97.2) |  |  |  |
| Non-White |  | 2 | 2 |  |  |  |
| **Education - Total (%)** |  |  |  | Fisher's exact test | 0.2512 |  |
| Highschool or Below |  | 12 (11.1) | 10 (13.9) |  |  |  |
| College |  | 53 (49.7) | 42 (58.3) |  |  |  |
| Graduate School |  | 43 (39.8) | 20 (27.8) |  |  |  |
| **Apolipoprotein E4 Carrier - Total (%)** |  | 35 (31.82) | 14 (19.44) | Fisher's exact test | 0.0871 |  |
| **Depression Measure** |  |  |  |  |  |  |
| **CES-D - Mean (SD) [Range]** |  |  |  |  |  |  |
| Total Score |  | 7.32 (5.73) [0-29] | 7.77 (5.46) [0-24] | Unpaired two-tailed t test | 0.5981 |  |
| **Sleep Measures** |  |  |  |  |  |  |
| **Self-Reported Sleep Duration - Mean (SD)** |  |  |  |  |  |  |
| Sleep Duration |  | 6.67 (0.87) | 6.90 (1.00) | Unpaired two-tailed t test | 0.1019 |  |
| Sleep Change (Slope) |  | -0.15 (0.28) | -0.23 (0.29) | Unpaired two-tailed t test | 0.0861 |  |
| Sleep Variability (SD) |  | 0.74 (0.32) | 0.80 (0.32) | Unpaired two-tailed t test | 0.2644 |  |
| **Sleep Phenotype - Total (%)** | | | | $\chi^2$ | 0.0608 | |
| Short Sleeper: Median <7 hrs. |  | 41 (38.0) | 22 (30.6) |  |  |  |
| Medium Sleeper: Median 7 hrs. |  | 50 (46.3) | 28 (38.9) |  |  |  |
| Long Sleeper: Median >7 hrs. |  | 17 (15.7) | 22 (30.6) |  |  |  |
| **Cognitive Measures** |  |  |  |  |  |  |
| **Cognitive Test Score - Mean (SD) [Range]** |  |  |  |  |  |  |
| Mini-Mental State Examination Score |  | 27.96 (2.59) [14-30] | 26.08 (3.46) [16-30] | Unpaired two-tailed t test | <0.0001 |  |
| Mattis Dementia Rating Scale |  | 139.6 (5.13) [112-144] | 133.4 (10.39) [75-144] | Unpaired two-tailed t test | <0.0001 |  |
| Trail Making Test B |  | 98.34 (65.74) [19-442] | 177.3 (98.31) [8-486] | Unpaired two-tailed t test | <0.0001 |  |
| WAIS - Vocabulary Age-Scaled |  | 12.93 (2.15) [7-19] | 11.96 (2.05) [7-19] | Unpaired two-tailed t test | 0.0031 |  |
| WAIS - Comprehension Age-Scaled |  | 12.72 (2.11) [7-18] | 11.55 (1.88) [6-15] | Unpaired two-tailed t test | 0.0002 |  |
| WAIS - Block Design Age-Scaled |  | 13.60 (2.83) [3-19] | 11.09 (3.12) [0-18] | Unpaired two-tailed t test | <0.0001 |  |
| WAIS - Digit-Symbol Age-Scaled |  | 13.07 (3.01) [1-17] | 9.72 (2.90) [0-15] | Unpaired two-tailed t test | <0.0001 |  |
| Cognitively Impaired (%) |  | 15 (30.6) | 34 (69.4) | Fisher's exact test | <0.0001 |  |

The association between sleep parameters and the time to cognitive impairment (reaching threshold on 2 or more cognitive tests) was assessed through a multivariate Cox proportional hazards regression model. To estimate hazard ratios (HR) and 95% confidence intervals (CI) multivariable adjusted Cox models were computed. Participants were removed prior to analysis if they had not completed at least 3 testing sessions (enabling analysis of standard deviation of sleep), reducing the sample size to n=553. In the multivariable analyses, **Model 1** controlled for demographic covariates of gender (female, male), ethnicity (white, non-white), years of education (at or below high school, college, graduate school), and Apolipoprotein Ɛ4 carrier status (carrier, non-carrier). **Model 2** included the demographic variables in **Model 1** with the addition of CES-D median depression score. Final **Model 3** added in sleep factors: change in sleep duration (“Sleep Change”), variability in sleep duration (“Sleep Variability”), and categorical assessment of sleep phenotype (“Short Sleeper”, “Long Sleeper”).

The ability of sleep parameters at baseline to predict incident cognitive impairment over an interval of 10 years was assessed using multivariate linear regression. For these analyses, we evaluated only participants with at least 3 Health Behavior Questionnaire assessments beginning at least 10 years prior to their final cognitive assessment. This resulted in a sample size of n=180 participants. In the multivariable analyses, **Model 1** controlled for demographic covariates gender (female, male), ethnicity (white, non-white), years of education (at or below high school, college, graduate school), and Apolipoprotein Ɛ4 carrier status (carrier, non-carrier). **Model 2** included the demographic variables in **Model 1** with the addition of CES-D median depression score. Final **Model 3** added in sleep factors: Sleep Variability, and categorical assessment of Short Sleeper and Long Sleeper phenotypes.

## Results

### Participant demographics and self-reported sleep parameters

A flow chart for study participant inclusion and exclusion is provided in **Figure 1**. We evaluated data from 1104 SLS participants. The SLS includes repeated neurocognitive assessment, self-report on the HBQ, and the CES-D test. From these initial 1104 participants, we excluded any lacking demographic data (n=282), primarily *APOE* genotype, leaving an initial study sample (**Sample 1**) of 822 participants. Demographic data for **Sample 1** is presented in **Table 1 (top)** with participants separated into three age brackets: <65 yrs, 65-84 yrs, and ≥85 yrs of age at final testing date. In this sample, there was a significant association between age and years of education, with those in the older age brackets having lower levels of educational attainment (χ^2^=21.09; df=4; p=0.0003). No significant difference in *APOE* allele status was observed between age groups.

Depression was assessed with the CES-D test. We calculated median CES-D score among all study visits for each participant and compared them among these age groups. Differences between age groups in global CES-D score were significant (F (2, 819)=5.625, p=0.0037; One-Way ANOVA), with 65–84-year-old participants reporting significantly lower levels of depression compared to participants under 65 (p=0.0076, Šídák’s multiple comparisons test).

The Health Behavior Questionnaire (HBQ) item #100 asks about average nightlysleep duration over the past week (not including naps). We calculated the median sleep duration among all study visits for each participant. We observed a significant difference in median sleep duration among the age groups (F (2, 819) = 9.432, p<0.0001; One-Way ANOVA), with longer sleep observed with increasing age (<65 yrs vs. 65-84 yrs, p=0.0035; <65 yrs vs. ≥85 yrs, p<0.0001; Šídák’s multiple comparisons test). We next characterized participant sleep duration categorically as “Short Sleepers”, “Medium Sleepers” and “Long Sleepers”. Initially, we defined Short Sleep as ≤6 hrs., Medium Sleep as 6-9 hrs., and Long Sleep as ≥9 hrs. However, with this breakdown, representation among the shortest and longest sleepers was minimal, particularly among participants ≥85 years of age. To achieve broader distribution, we classified “Short Sleepers” as those with a median of <7 hours per night, “Medium Sleepers” as those with a median of 7 hours per night, “Long Sleepers” as those with a median of >7 hours per night. Within this study population, this approach resulted in roughly equal participant distribution among these three categories. We observed significant differences in the distribution of these classifications among age groups (χ^2^=20.30; df=4; p=0.0004), with an increasing percentage of “Long Sleepers” and fewer “Short” and “Medium Sleepers” with advancing age.

The SLS evaluated cognitive function using four tests: the MMSE, the Mattis dementia rating scale, Trails B, and WAIS. As shown in **Table 1 (top), Sample 1** expressed a significant decrease in cognitive performance across all tests with age. As expected, there was a higher percentage of cognitive impairment expressed in participants as they aged (χ^2^=105.6; df=2; p<0.0001). Of the 822 participants, 196 participants (23.8%) presented with cognitive impairment during their final cognitive evaluation.

### Sleep Duration and longitudinal Sleep Variability are associated with cognitive impairment

We next evaluated the relationship between longitudinal sleep parameters and cognitive impairment using survival analysis with Cox Proportional Hazard Regression. For this analysis, we excluded participants from **Sample 1** that did not have at least three HBQ (which includes sleep duration) assessments (n=269), resulting in a sub-sample (**Sample 2**) of n=553 participants (**Figure 1**). **Table 1 (middle)** provides the same demographic outputs as **Sample 1**, but for **Sample 2**. Similar to **Sample 1**, **Sample 2** had significantly longer median sleep duration within the ≥85-year-old group (F (2, 550)=6.225, p=0.0018; One-Way ANOVA; <65 vs. ≥85 p=0.0012, Šídák’s multiple comparisons test) and a similar shift in sleep phenotype distribution towards “Longer Sleepers” with age (χ^2^=13.82; df=4; p=0.0079). Additionally, the inclusion of at least 3 HBQ allowed for evaluation of both the Sleep Change and Sleep Variability within **Sample 2**. We did not observe any effect of age group on the Sleep Change or Sleep Variability. Within **Sample 2**, as with **Sample 1**, there was a significantly increase in cognitive impairment with age (χ^2^=57.30; df=2; p<0.0001), with 94 (17.0%) out of 553 reaching threshold for cognitive impairment by their last cognitive assessment.

For the survival analysis, we included sleep and depression data up to and including the visit where participants ‘converted’ to the status of cognitively impaired (having 2 or more cognitive measures below age-normed thresholds). For participants that remained cognitively intact for the duration of the study, all data were included. In our initial model (**Model 1**), we evaluated the contribution of age, gender, years of education, ethnicity and *APOE* status on the incidence of cognitive impairment over time using a Cox proportional hazard regression approach. **Model 1** output is shown in **Table 2** (top). Within this model possessing one or more copies of the *APOE4* allele was significantly associated with cognitive impairment (HR 1.98, 95% CI 1.26, 3.09). We next developed a second model (**Model 2**; **Table 2**, middle) which added the contribution of depression (median CES-D total score) to **Model 1**. Within this model, both *APOE4* status (HR 2.08, 95% CI 1.33, 3.25) and higher depression score (HR 1.06, 95% CI 1.02, 1.10) were significantly associated with a higher risk of cognitive impairment. **Model 1** had concordance value of 0.62 while **Model 2** had a concordance value of 0.68, suggesting that the inclusion of depression status is informative in describing the contributors to cognitive impairment in this population.

**Table 2.** Cox proportional hazard regression models evaluating longitudinal associations with cognitive impairment. HR: Hazard Ratio, CI: Confidence Interval. CES-D: Center for Epidemiological Studies Depression Scale; SD: Standard Deviation.

| Cox Proportional Hazard Model Survival Analysis (Sample 2, n = 553) |  |  |  |
| --- | --- | --- | --- |
| Model | Covariates | HR (95% CI) | p value |
| Model 1<br>Demographic Variables<br>**Concordance: 0.62** | Gender | 1.35 (0.87-2.09) | 0.18 |
|  | Ethnicity | 0.49 (0.21-1.15) | 0.10 |
|  | Years of Education | 1.06 (0.98-1.14) | 0.12 |
|  | **Apolipoprotein E4 carrier** | 1.98 (1.26-3.09) | **<0.005** |
| Model 2<br>Demographic Variables<br>+<br>Depression<br>**Concordance: 0.68** | Gender | 1.46 (0.93-2.28) | 0.10 |
|  | Ethnicity | 0.49 (0.21-1.16) | 0.11 |
|  | Years of Education | 1.07 (0.99-1.16) | 0.07 |
|  | **Apolipoprotein E4 carrier** | 2.08 (1.33-3.25) | **<0.005** |
|  | **Depression (median CES-D total score)** | 1.06 (1.02-1.10) | **<0.005** |
| Model 3<br>Demographic Variables<br>+<br>Depression<br>+<br>Sleep Factors<br>**Concordance: 0.70** | **Gender** | 1.69 (1.07-2.68) | **0.03** |
|  | Ethnicity | 0.58 (0.24-1.40) | 0.23 |
|  | **Years of Education** | 1.11 (1.02-1.20) | **0.01** |
|  | **Apolipoprotein E4 carrier** | 2.36 (1.49-3.74) | **<0.005** |
|  | **Depression (median CES-D total score)** | 1.06 (1.02-1.09) | **<0.005** |
|  | Short Sleeper | 1.61 (0.96-2.69) | 0.07 |
|  | Long Sleeper | 1.24 (0.73-2.12) | 0.43 |
|  | Sleep Change (slope) | 2.38 (0.14-41.69) | 0.55 |
|  | **Sleep Variability (SD)** | 2.21 (1.27-3.86) | **0.01** |

In our third model (**Model 3**), we added three longitudinal sleep parameters to **Model 2**. This included categorical sleep duration phenotype (Short, Medium, and Long Sleeper), Sleep Change, and Sleep Variability. **Model 3** (**Table 2**, bottom) had a concordance of 0.70, suggesting that the inclusion of longitudinal sleep parameters better described the risk for cognitive impairment within this study population. In this model, as in **Model 2**, we observed that being female (HR 1.69, 95% CI 1.07, 2.68), years of education (HR 1.11, 95% CI 1.02, 1.20), *APOE4* status (HR 2.36, 95% CI 1.49, 3.74), and higher depression scores (HR 1.05, 95% CI 1.02, 1.09) were significantly associated with risk of cognitive impairment. In addition, having higher Sleep Variability (HR 2.47, 95% CI 1.35, 4.54) was significantly associated with cognitive impairment. Additionally, expressing a “Short Sleep” phenotype exhibited a trend towards an association with cognitive impairment (p=0.07; HR 1.61, 95% CI 0.96, 2.69), while neither categorization as a “Long Sleeper”, nor Sleep Change contributed to cognitive impairment in this model.

A forest plot depicting the log hazard ratios for each **Model 3** covariate is shown in **Figure 2A**. **Figures 2B-D** present the survival curves depicting the relationships between gender (**Figure 2B**), years of education (**Figure 2C**), *APOE4* allele status (**Figure 2D**), depression (**Figure 2E)**, Short Sleeper phenotype (**Figure 2F**), and Sleep Variability (**Figure 2G**), age and ‘survival’ with intact cognition versus cognitive impairment. Increased risk of cognitive impairment is reflected in the leftward shift in the survival curve with female gender, fewer years of educational attainment, *APOE4* allele status, higher depression indicated by median CES-D total score, Short Sleeper phenotype, and higher Sleep Variability.

**Figure 2.**
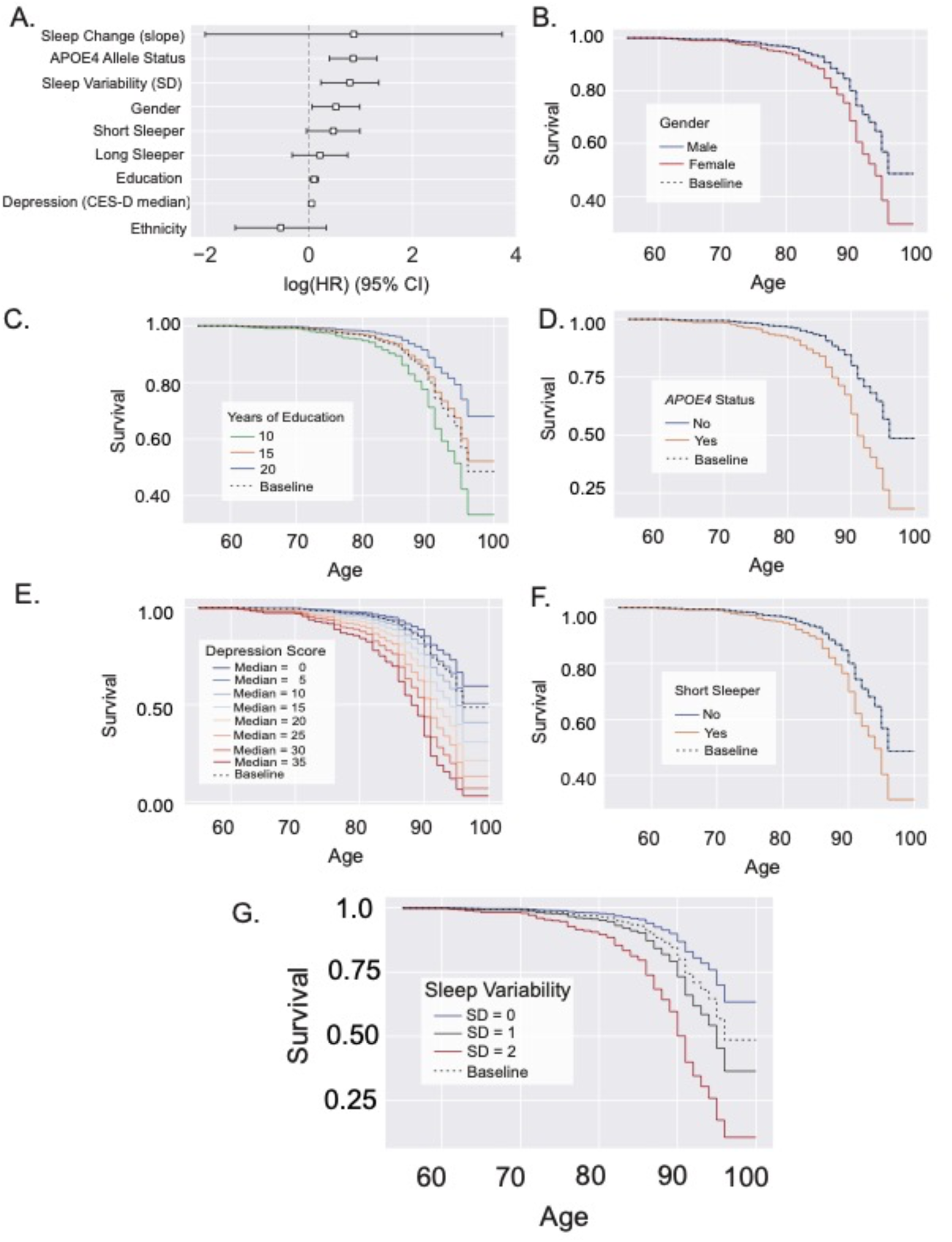
Short sleep duration and sleep variability are associated with cognitive impairment. **(A)** Forest plot displaying hazard ratios of all variables evaluated within Model 3. **(B-D)** Female gender **(B)**, lower levels of education (**C**), the presence of at least one APOE4 allele **(D)**, and higher scores on the CES-D depression scale **(E)** are associated with higher risk for cognitive impairment. Classification as a “Short Sleeper” (<7 hrs.) (**F)** and higher levels of Sleep Variability **(G)** are associated with greater risk of cognitive impairment. CES-D: Center for Epidemiological Studies Depression Scale; SD: Standard Deviation.

### Sleep variability predicts cognitive impairment 10-years later

In agreement with prior studies, our analysis showed that positive *APOE4* allele status, fewer years of education, and depression are significantly associated with cognitive impairment. They further showed that the increased Sleep Variability and perhaps Short Sleep phenotype are associated with age-related cognitive impairment.

In our final analysis, we took advantage of this long-term longitudinal dataset to test whether these factors at baseline predicted the incident development of cognitive impairment over an interval of 10 years. For these analyses, we evaluated only participants with at least 3 Health Behavior Questionnaire assessments beginning at least 10 years prior to their final cognitive assessment. This resulted in a sample size of n=180 participants. **Table 1 (bottom)** provides the demographic information for this, “**Sample 3**”. Because inclusion in this analysis required many years of longitudinal data collection, few participants were present in the <65 age range. Thus, for this analysis, we binned participants into two age groups: <85, and ≥85 years of age. We conducted three multivariate linear regression prediction analyses using a similar model structure as above. Model outputs are provided in **Table 3**. In **Model 1’**, only age (β=0.0545; p<0.0001) was a significant predictor of cognitive impairment at a 10-year interval. In **Model 2’**, both age (β=0.0586; p<0.0001) and depression (β=0.0339; p=0.003) were significant predictors of cognitive impairment at a 10-year interval. In **Model 3’**, age (β=0.0607; p<0.0001), depression (β = 0.0262; p=0.025) and Sleep Variability (β=0.3491; p=0.048) were significant predictors of cognitive impairment over a 10-year period. Based on adjusted R^2^ values, (**Model 1’** R^2^=0.141, F (5, 174)=6.888, p=6.84E-06; **Model 2’** R^2^=0.178, F (6, 173)=7.469, p=3.97E-07; **Model 3’** R^2^=0.201, F (9, 170)=6.010, p=2.67E-07), each successive model better accounted for prospective risk of cognitive impairment within this study population.

**Table 3.**
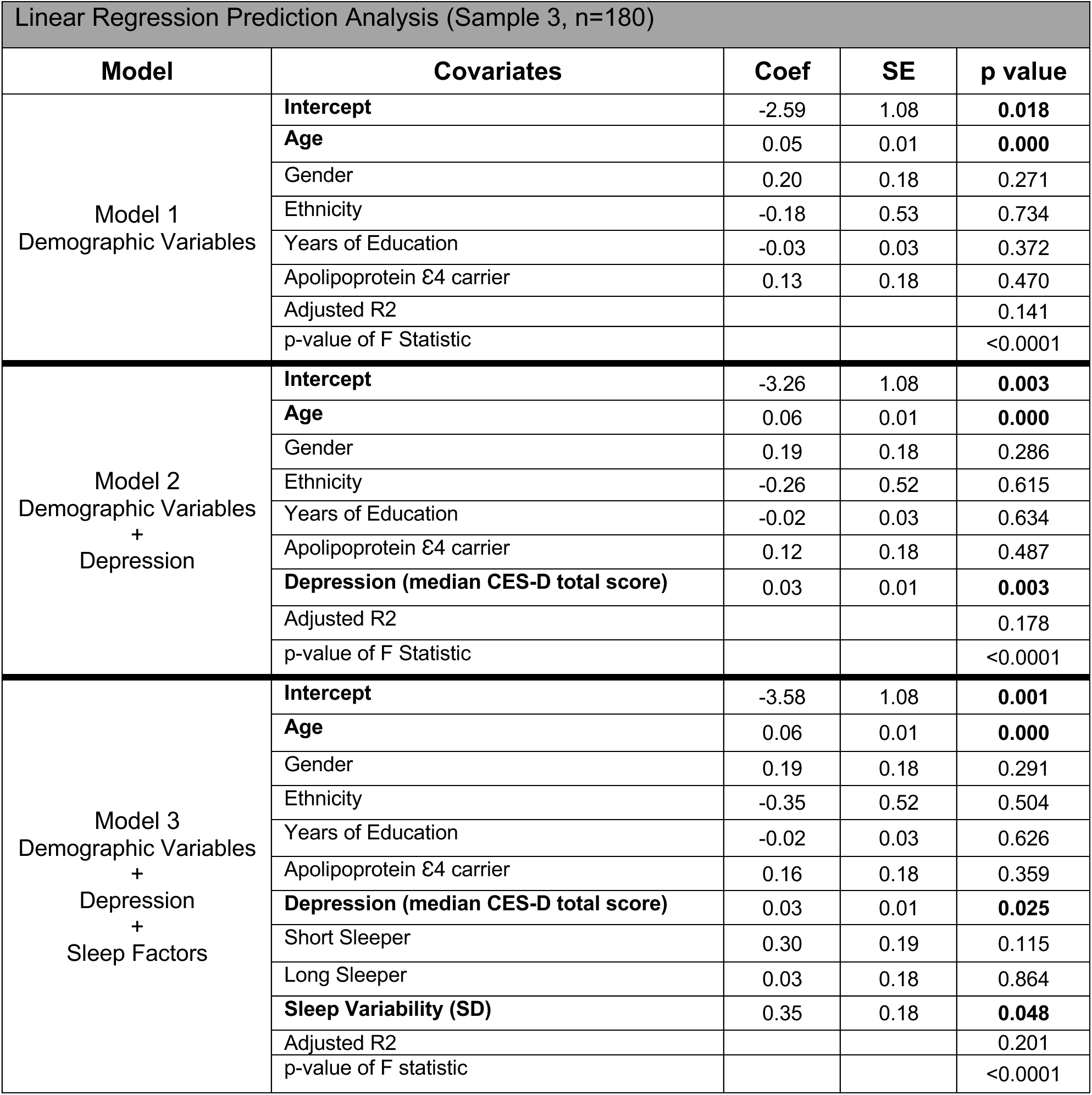
Multivariate linear regression models predicting incident cognitive impairment. HR: Hazard Ratio; CI: confidence interval; CES-D: Center for Epidemiological Studies Depression Scale; SD: Standard Deviation.

## Discussion

Analysis of longitudinal measures of self-reported sleep duration within the community-based SLS sample demonstrated for the first timethat variability in sleep duration across measurements was not only significantly associated with cognitive impairment, but also predicted the development of cognitive decline over a 10-year period. Additionally, consistent with prior studies [10–13] we observed a trend towards an association between “Short Sleep” (<7 hrs per night) and cognitive impairment.

The relationship between short sleep and risk of cognitive impairment has previously been observed between in healthy aging adults [10–13]. There are several potential mechanistic explanations for this clinical association. Chronic sleep disruption is associated with cardio/cerebrovascular disease [32], stroke [33, 34], metabolic syndrome/diabetes [35], and depression [36], each of which may contribute to the observed increased risk of cognitive decline [37–40]. Short sleep duration has also been associated with greater amyloid β (Aβ) plaque burden [15] and faster ventricular expansion [10], supporting notion that short sleep duration exacerbates AD-related neurodegenerative processes. Studies in rodents demonstrate that sleep-active glymphatic function contributes to the clearance of Aβ, tau and α-synuclein, while acute sleep deprivation slows this clearance. A recent study utilizing intrathecal contrast-enhanced MRI confirmed that solute clearance from the human brain was sleep dependent[41]. Thus the chronic impairment of sleep-active glymphatic function may underlie the observed clinical association of short sleep duration and cognitive impairment.

The SLS provides the unique opportunity for assessing longitudinal changes in self-reported sleep duration, allowing for evaluation of both the change in sleep duration (slope), and variability in that self-report (standard deviation) over decades of assessment. Our study did not find, as previous studies did [42, 43], that an increase or decrease in sleep duration was associated with incidence of cognitive impairment. However, this study is the first to report that variability in sleep duration was both significantly associated and predictive of impaired cognitive function.

This increased variability reflects a widening range in sleep durations over the course of many years. Yet it remains unclear exactly what such variability reflects. Assuming that self-reported sleep duration aligns closely with objective sleep duration, an assumption that may not be completely valid, variability in this study may reflect age-related changes sleep architecture and duration in the aging brain [44]. However, the absence of an association with the overall slope in sleep duration argues against this possibility. It is possible that the observed variability reflects changes only in self-reported, and not objective sleep duration over time. Such changes may originate from cognitive and psychological influences [45]. It is also possible that this variability relates to comorbid neurological or psychiatric changes with age, such as depression, chronic pain, or frequent nocturia, or to changes in social and behavioral factors [46], such as changes in shift work, retirement or changes in marital status. Future studies will need to define the relationship between long-term variability in self-reported and objective sleep duration, and their associations with these cognitive, psychological, social and behavioral factors.

Importantly, our study also highlights how different experimental approaches to defining longitudinal changes in sleep patterns may result in widely varying results. Discrepancies in methodological approach, including approaches to assessing sleep duration and quality (e.g., self-report, actigraphy, EEG), frequency and duration of assessment (single time point assessment, multi-day, long-term ecological assessments), study timescales (from days to years), and synchrony of data collection with cognitive and other assessments, can each substantially alter study findings. Thus, this study supports the growing need for a more comprehensive evaluation of longitudinal sleep behavior in future studies.

### Strengths and Limitations

The overarching goal of the SLS, beginning in 1956, was to identify the combined impact of aging and birth cohorts with health and lifestyle factors contributing to downstream cognitive impairment [16]. While this study did not evaluate birth cohorts, due to the breadth of study duration, the SLS provides a unique opportunity to evaluate both self-reported sleep duration changes and cognitive performance in nearly 1100 participants assessed over decades. As a retrospective analysis of SLS data, in the present study, sleep duration was defined only via self-report in a single question within the HBQ. While this permits the assessment of longitudinal changes in sleep patterns, as discussed, the subjective natures of the measure and the infrequent sampling limit the conclusions that we can draw from these data.

Similarly, the present study does not offer the opportunity to assess clinical sleep disorders, such as obstructive sleep apnea [47, 48] or insomnia [49], which have previously been associated with cognitive impairment. Such conditions are frequently present years prior to symptoms of cognitive decline [7, 8], and may be contributing to unstable sleep durations among the participants of this study.It is also important to note that these sleep disorders are often co-morbid with depression. Along these lines, because longitudinal data on cognitive performance, depression (measured by CES-D) and sleep duration were available throughout SLS, we were able to demonstrate a distinct relationship between age, variability in sleep duration, and depression in predicting incident cognitive decline.

## Conclusions

The present study demonstrates that in addition to short sleep duration, instability in longitudinal self-reported sleep duration is associated with cognitive impairment in the community-based SLS sample. Further, it demonstrates that this longitudinal variability in sleep duration predicted incident cognitive impairment over a 10-year interval. These findings suggest that longitudinal changes in sleep behavior, rather than sleep duration alone, may be important contributors to the development of age-related cognitive impairment. They argue that to understand the clinical relationship between age, sleep disruption, and cognitive impairment, it may be necessary to assess changes in sleep behavior over longer time scales than is presently in common research practice.

## Data Availability

All data produced in the present work are a part of the collective Seattle Longitudinal Study

## Acknowledgements

P30AG066518 to LCS, MML; RX002947 to JEE.; AG052354 SAK; AG055653-03 SW; AG054456 JJI.

## References

1. Collaborators, G.B.D.D.F., Estimation of the global prevalence of dementia in 2019 and forecasted prevalence in 2050: an analysis for the Global Burden of Disease Study 2019. Lancet Public Health, 2022. 7(2): p. e105–e125.

2. 2021 Alzheimer’s disease facts and figures. Alzheimers Dement, 2021. 17(3): p. 327–406.

3. Brookmeyer, R., et al., Forecasting the global burden of Alzheimer’s disease. Alzheimers Dement, 2007. 3(3): p. 186–91.

4. Villemagne, V.L., et al., Amyloid beta deposition, neurodegeneration, and cognitive decline in sporadic Alzheimer’s disease: a prospective cohort study. Lancet Neurol, 2013. 12(4): p. 357–67.

5. Holdridge, K.C., et al., Targeting amyloid beta in Alzheimer’s disease: Meta-analysis of low-dose solanezumab in Alzheimer’s disease with mild dementia studies. Alzheimers Dement, 2023.

6. van Dyck, C.H., et al., Lecanemab in Early Alzheimer’s Disease. N Engl J Med, 2023. 388(1): p. 9–21.

7. Sindi, S., et al., Sleep disturbances and dementia risk: A multicenter study. Alzheimers Dement, 2018. 14(10): p. 1235–1242.

8. Peter-Derex, L., et al., Sleep and Alzheimer’s disease. Sleep Med Rev, 2015. 19: p. 29–38.

9. Holth, J., T. Patel, and D.M. Holtzman, Sleep in Alzheimer’s Disease - Beyond Amyloid. Neurobiol Sleep Circadian Rhythms, 2017. 2: p. 4–14.

10. Lo, J.C., et al., Sleep duration and age-related changes in brain structure and cognitive performance. Sleep, 2014. 37(7): p. 1171–8.

11. Chen, J.C., et al., Sleep duration, cognitive decline, and dementia risk in older women. Alzheimers Dement, 2016. 12(1): p. 21–33.

12. Keage, H.A., et al., What sleep characteristics predict cognitive decline in the elderly? Sleep Med, 2012. 13(7): p. 886–92.

13. Tworoger, S.S., et al., The association of self-reported sleep duration, difficulty sleeping, and snoring with cognitive function in older women. Alzheimer Dis Assoc Disord, 2006. 20(1): p. 41–8.

14. Sprecher, K.E., et al., Poor sleep is associated with CSF biomarkers of amyloid pathology in cognitively normal adults. Neurology, 2017. 89(5): p. 445–453.

15. Spira, A.P., et al., Self-reported sleep and beta-amyloid deposition in community-dwelling older adults. JAMA Neurol, 2013. 70(12): p. 1537–43.

16. Schaie, K.W., Developmental influences on adult intelligence : the seattle longitudinal study. 2nd ed. 2013, New York, NY: Oxford University Press. viii, 587 pages.

17. Schaie, K.W., et al., Extending neuropsychological assessments into the primary mental ability space. Neuropsychol Dev Cogn B Aging Neuropsychol Cogn, 2005. 12(3): p. 245–77.

18. Boron, J.B., S.L. Willis, and K.W. Schaie, Cognitive training gain as a predictor of mental status. J Gerontol B Psychol Sci Soc Sci, 2007. 62(1): p. P45–52.

19. Schaie, K.W., S.L. Willis, and A.M. O’Hanlon, Perceived intellectual performance change over seven years. J Gerontol, 1994. 49(3): p. P108–18.

20. Hofer, S.M. and D.F. Alwin, Handbook of cognitive aging : interdisciplinary perspectives; Part VII: Chapter 39 Midlife Cognition: The Association of Personality With Cognition and Risk of Cognitive Impairment. 2008, Los Angeles: Sage Publications. xiii, 730 pages.

21. Vertesi, A., et al., Standardized Mini-Mental State Examination. Use and interpretation. Can Fam Physician, 2001. 47: p. 2018–23.

22. Zhang, S., et al., Determining Appropriate Screening Tools and Cutoffs for Cognitive Impairment in the Chinese Elderly. Front Psychiatry, 2021. 12: p. 773281.

23. Boban, M., et al., The reliability and validity of the mini-mental state examination in the elderly Croatian population. Dement Geriatr Cogn Disord, 2012. 33(6): p. 385–92.

24. Mattis, S., Dementia rating scale: professional manual. 1988: Psychological Assessment Resources, Incorporated.

25. Paolo, A.M., et al., Differentiation of the dementias of Alzheimer’s and Parkinson’s disease with the dementia rating scale. J Geriatr Psychiatry Neurol, 1995. 8(3): p. 184–8.

26. Lukatela, K.A., et al., Dementia rating scale performance: a comparison of vascular and Alzheimer’s dementia. J Clin Exp Neuropsychol, 2000. 22(4): p. 445–54.

27. Tombaugh, T.N., Trail Making Test A and B: normative data stratified by age and education. Arch Clin Neuropsychol, 2004. 19(2): p. 203–14.

28. Crum, R.M., et al., Population-based norms for the Mini-Mental State Examination by age and educational level. JAMA, 1993. 269(18): p. 2386–91.

29. Spreen, O. and E. Strauss, A compendium of neuropsychological tests : administration, norms, and commentary. 1991, New York: Oxford University Press. xv, 442 p.

30. Lewinsohn, P.M., et al., Center for Epidemiologic Studies Depression Scale (CES-D) as a screening instrument for depression among community-residing older adults. Psychol Aging, 1997. 12(2): p. 277–87.

31. Radloff, L.S., The CES-D Scale: a self-report depression scale for research in the general population. Vol. 1. 1977: Applied Psychological Measurement.

32. Ferrie, J.E., et al., A prospective study of change in sleep duration: associations with mortality in the Whitehall II cohort. Sleep, 2007. 30(12): p. 1659–66.

33. Khot, S.P. and L.B. Morgenstern, Sleep and Stroke. Stroke, 2019. 50(6): p. 1612–1617.

34. Cai, H., X.P. Wang, and G.Y. Yang, Sleep Disorders in Stroke: An Update on Management. Aging Dis, 2021. 12(2): p. 570–585.

35. Grandner, M.A., et al., Sleep Duration and Diabetes Risk: Population Trends and Potential Mechanisms. Curr Diab Rep, 2016. 16(11): p. 106.

36. Tsuno, N., A. Besset, and K. Ritchie, Sleep and depression. J Clin Psychiatry, 2005. 66(10): p. 1254–69.

37. Verdelho, A., et al., Cognitive impairment in patients with cerebrovascular disease: A white paper from the links between stroke ESO Dementia Committee. Eur Stroke J, 2021. 6(1): p. 5–17.

38. Al-Qazzaz, N.K., et al., Cognitive impairment and memory dysfunction after a stroke diagnosis: a post-stroke memory assessment. Neuropsychiatr Dis Treat, 2014. 10: p. 1677–91.

39. Ehtewish, H., A. Arredouani, and O. El-Agnaf, Diagnostic, Prognostic, and Mechanistic Biomarkers of Diabetes Mellitus-Associated Cognitive Decline. Int J Mol Sci, 2022. 23(11).

40. Chakrabarty, T., G. Hadjipavlou, and R.W. Lam, Cognitive Dysfunction in Major Depressive Disorder: Assessment, Impact, and Management. Focus (Am Psychiatr Publ), 2016. 14(2): p. 194–206.

41. Lee, S., et al., Contrast-enhanced MRI T1 Mapping for Quantitative Evaluation of Putative Dynamic Glymphatic Activity in the Human Brain in Sleep-Wake States. Radiology, 2021. 300(3): p. 661–668.

42. Ferrie, J.E., et al., Change in sleep duration and cognitive function: findings from the Whitehall II Study. Sleep, 2011. 34(5): p. 565–73.

43. Devore, E.E., et al., Sleep duration in midlife and later life in relation to cognition. J Am Geriatr Soc, 2014. 62(6): p. 1073–81.

44. Schmidt, C., P. Peigneux, and C. Cajochen, Age-related changes in sleep and circadian rhythms: impact on cognitive performance and underlying neuroanatomical networks. Front Neurol, 2012. 3: p. 118.

45. Althubaiti, A., Information bias in health research: definition, pitfalls, and adjustment methods. J Multidiscip Healthc, 2016. 9: p. 211–7.

46. Bliwise, D.L. and T.B. Young, The parable of parabola: what the U-shaped curve can and cannot tell us about sleep. Sleep, 2007. 30(12): p. 1614–5.

47. Yaffe, K., et al., Sleep-disordered breathing, hypoxia, and risk of mild cognitive impairment and dementia in older women. JAMA, 2011. 306(6): p. 613–9.

48. Osorio, R.S., et al., Sleep-disordered breathing advances cognitive decline in the elderly. Neurology, 2015. 84(19): p. 1964–71.

49. Cricco, M., E.M. Simonsick, and D.J. Foley, The impact of insomnia on cognitive functioning in older adults. J Am Geriatr Soc, 2001. 49(9): p. 1185–9.

